# Association between depressive symptoms and domestic violence and abuse among Nepali Women: A cross-sectional study

**DOI:** 10.1101/2025.09.17.25335959

**Authors:** Bipsana Shrestha, Yunika Acharya, Priyanka Timsina, Pratiksha Paudel, Aerona Karmacharya, Soniya Makaju, Bandana Paneru, Archana Shrestha

## Abstract

**Background:** Domestic violence is a widespread public health problem and serious violation of human’s right. It occurs worldwide, across all generations, nationalities, communities and spheres of societies, irrespective of age, ethnicity, disability or other background. While men may also fall victim to domestic violence, it is women who bear the more serious implications for their health. Domestic violence victims have a higher risk of post-traumatic stress disorders, and suicide. Despite a critical link between domestic violence and mental health, very few studies have been done to identify the link between violence and mental health problems in Nepal. Thus, to bridge the gap this study aims to investigate the association between depressive symptoms and different forms of domestic violence; physical, sexual and emotional violence.

**Methods:** We conducted a cross-sectional study among 429 women aged 30 to 60 years who underwent cervical cancer screening in Dhulikhel and Banepa municipalities, Kavrepalanchowk district, Nepal. We conducted the study amid the COVID-19 pandemic. Therefore, interviews were conducted over the phone for infection prevention. We used univariable and multivariable logistic regression models to assess the association between depressive symptoms and domestic violence and abuse.

**Results:** Findings of our study suggest 2.99 times (95% CI: 1.57, 5.67, p = 0.001) higher odds of having depressive symptoms in women experiencing any type of violence. Similarly, the odds of having depressive symptoms were 2.5 times (95% CI: 1.26, 4.98, p=0.009) higher in participants who had experienced physical violence and 6.21 times (95% CI: 2.49, 15.46, p<0.001) higher among the participants who had experienced sexual violence or emotional abuse.

**Conclusion:** Those women who have experienced physical violence, sexual abuse, or emotional abuse are more likely to experience depressive symptoms. To better understand the factors contributing to these symptoms, further in-depth and exploratory studies are needed. Community-based interventions are needed to offer counseling, support groups, and safe spaces for women affected by violence.

## Background

Domestic violence is a widespread public health problem and serious violation of human’s right (1). It occurs worldwide, across all generations, nationalities, communities and spheres of societies, irrespective of age, ethnicity, disability or other background (2). Domestic violence can manifest in various forms including: verbal (shouting, blaming, taunting, conflicts), physical (beating, shoving, pushing, using hands, legs, sticks like means), emotional (feeling anxious or depressed due to conflicts with in-laws or husband), or sexual (unwanted sexual acts, comments, sexual coercion) (3,4). Despite formulation of international and national laws and regulations to prevent violence, domestic violence is a problem deeply rooted within the realm of gender inequality, patriarchy, constructions of masculinities and femininities which act to normalize violence between the sexes (5).

Studies from 161 countries indicate that around 30% of women have ever experienced violence of any kind, be it from a current or a former partner or from a non-partner (6). In Nepal – a country characterized by the dominant patriarchal systems accompanied by traditional gender roles – the state of affairs is gravely concerning (7). According to the latest Nepal Demographic and Health Survey (NDHS), 23% of women aged 15-49 reported experiencing physical violence, 7% reported having experienced sexual violence, and 13% reported emotional violence during their lifetime (8). These estimates might be underestimated as reporting rates of domestic violence in Nepal tend to be very low. This is because most women hesitate to admit to any abuse for fear of being stigmatized, a lack of faith in support systems or protection, and in many cases because of threats from the abuser (9,10).

A systematic review of domestic violence risk factors have highlighted the multifaceted nature of violence, emphasizing the interplay between individual, familial, and societal influences (11). Many women experience physical violence, often at the hands of their husbands who are under the influence of alcohol (12). Women who lack decision-making autonomy and have controlling husbands were at greater risk of intimate partner violence (IPV) (13). Women in Nepal often rely on their husbands for financial support which is found to be one of the major reasons for IPV in Nepal (14). The control exerted by husbands or in-laws can lead to constant emotional distress. This emotional violence includes verbal abuse, humiliation, and constant threats, all of which severely affect women’s mental health (15). Sexual violence is another prevalent form of abuse in Nepal, though it remains largely unreported due to societal stigma and legal barriers (16). Many women in abusive marriages experience forced sex, which is often normalized or minimized by societal beliefs that view a wife’s consent as unnecessary within marriage (16).

Despite substantial evidence on the health consequences of gender-based violence (GBV) globally, Nepal’s health sector has been slow to recognize GBV as a legitimate public health problem. It was only in 2010 after development of the National Plan of Action against GBV, GBV was incorporated as a health sector response (17). Over the past 20 years, Nepal has made significant strides towards ensuring gender equality and ending GBV through inclusion in Three year plan (2010–2013) with an objective to eliminate GBV and promote gender equality and women’s empowerment (17). The National Safe Motherhood Plan (2002–2017) recognizes GBV as an important issue in women’s health. In addition to this, the Ministry of Health has also set up One Stop Crisis Management Centers (OCMC) in all the states to assist the GBV victims (17). Despite the existence of legal provisions and policies aimed at addressing gender-based violence in Nepal, societal stigma remains a significant barrier to women seeking help or leaving abusive relationships. Many women hold the belief that it is justified for husbands to beat their wives, which leads to silence and isolation regarding their experiences (18). This deeply rooted societal acceptance of violence, combined with a lack of accessible support services, often discourages women from seeking assistance or leaving violent situations (18).

While men may also fall victim to domestic violence, it is women who bear the more serious implications for their health (1). According to World Health Organization (WHO) global and regional estimates of women experiencing IPV, the odds of major depression was 1.97 times higher and alcohol use disorders 1.82 times higher compared to women who did not experience violence whereas the risk of suicide among women was higher by 4.54 times (3). A scoping review conducted among women in the Southeast Asian region showed that those who experienced domestic violence face a significantly higher risk of suicidal behavior and psychological distress (19). The study found that women who experienced intimate partner violence reported significantly higher levels of depression and anxiety compared to those who did not (19). Likewise, a study conducted in Srilanka found that women exposed to IPV had a higher likelihood of engaging in self-harm and suicide attempts with adjusted odds ratios (AOR) indicating that physical/sexual abuse was associated with an AOR of 5.17, whereas psychological abuse had an AOR of 4.64 for self-harm (20). In Nepal, research shows that women who died by suicide often faced physical abuse shortly before their deaths contributing factors included marital conflicts, financial hardships, and mental health challenges like depression (21). In certain instances, survivors of domestic violence may blame themselves for their partner’s verbal abuse which leads to difficulty in forming future relationships (22).

Children who grow up in violent households are also at risk, often developing emotional and behavioral disturbances, and are more likely to experience or perpetrate violence later in life (1). While existing studies globally and in Nepal have identified domestic violence as a significant contributor to mental health issues such as depression, many fail to explore the specific associations between different forms of violence—physical, sexual, and emotional—and their distinct effects on mental health (23). Thus, to bridge the gap this study aims to investigate the association between depressive symptoms and different forms of domestic violence; physical, sexual and emotional violence. By examining the prevalence and severity of depressive symptoms among women experiencing domestic violence, we hope to inform future interventions and policies and develop culturally sensitive and effective responses to both domestic violence and the mental health crises it triggers.

## Methods

### Aim, design and setting

This study aims to investigate the association between depressive symptoms and different forms of domestic violence; physical, sexual and emotional violence. We conducted a cross-sectional study among women aged 30 to 60 years who registered for HPV self-sampling, offered by Dhulikhel Hospital in Dhulikhel and Banepa municipalities, Kavrepalanchowk district, Nepal. The data was collected between March 15 and June 15, 2021.

### Study participants and recruitment

We enrolled 429 women conveniently, of age group 30 years and above out of a total 1856 women participating in the cervical cancer screening program of Dhulikhel hospital. The sample size was estimated with 50% proportion of depression among women experiencing domestic violence (24), at 95% confidence interval and 5% allowable error.

The inclusion criteria of the study included: (1) Women 30 to 60 years; (2) Have intact uterus (3) Residents of Dhulikhel and Banepa. The exclusion criteria were: (1) Pregnant; (2) Previous history of cervical intraepithelial neoplasia (CIN) or cancer. (3) Having severe mental health conditions (4) having a hearing impairment and not being able to provide informed consent.

Our Research Assistants (RAs) contacted female community health volunteers (FCHV) and oriented them to the study objective and expectations. These FCHVs have worked in Nepal at the community level for a long time and play a pivotal role in the Nepali community health workforce, specializing in health education, counseling, outreach, and resource distribution. The FCHVs disseminated information about the study to women in their network and shared the contact details of the interested participants with the research assistants. Subsequently, the research assistants contacted the women by phone, outlining the study’s objectives and explaining their potential role. Women expressing interest were then formally enrolled in the study after verbal informed consent was obtained, ensuring the anonymity and confidentiality of their information.

### Data collection

The research assistants (RAs) had telephonic interviews with the participants using a standardized electronic questionnaire on kobotoolbox. We conducted the study amid the COVID-19 pandemic. Therefore, interviews were conducted over the phone for infection prevention.

### Study metrics

We used a questionnaire to assess socioeconomic characteristics, including age, marital status, education, ethnicity, and presence of domestic violence and presence of depressive symptoms.

### Domestic violence and abuse

Domestic violence and abuse was measured using a Nepali-translated version of the domestic violence module developed by USAID (25). The instrument consists of seven questions related to experiences of physical violence, three questions on sexual violence, and three questions on emotional abuse. All questions were presented as binary (yes/no) items. A respondent was classified as having experienced domestic violence if they answered “yes” to any of the questions. Respondents were categorized as having experienced physical, sexual, or emotional violence if they answered “yes” to any of the questions in the respective sections on physical, sexual, or emotional abuse.

### Depressive symptoms

We assessed depressive symptoms using the nepali translated 9-item Patient Health Questionnaire (PHQ-9) (26), which is based on the Diagnostic and Statistical Manual of Mental Disorders (DSM-IV) (27). Participants were asked to rate nine common symptoms of depression they experienced over the past two weeks, using a four-point Likert scale ranging from 0 (not present at all) to 3 (almost every day), yielding a total score between 0 and 27. A score of 10 or higher was used as the cut-off to indicate the presence of depressive symptoms (28).

### Data analysis

We summarized study participants characteristics using frequency and percentage for categorical; and means and standard deviation for numeric variables. We used univariable and multivariable logistic regression models to assess the association between depressive symptoms and domestic violence and abuse.

We calculated the prevalence of depression and domestic violence with 95% CI. In the multivariable model, we adjusted for the confounding variables. We reported crude and adjusted odds ratio with 95% confidence interval and p-value. All analyses were conducted using STATA version 13.0 (Stata Corp., College Station, Texas, USA) for cleaning, coding and statistical analysis.

## Results

Table 1 illustrates the socio-demographic information of the participants. The mean age of the participants was 41.97 (SD 8.07) years. Most of the participants (42.9%) belonged to the age group 30-39 years and the majority of the participants were married (93%).

**Table 1.** Sociodemographic characteristics (n=429).

| **Sociodemographic characteristics** | **Frequency** | **Percentage (%)** |
| --- | --- | --- |
| **Age (mean±SD)** | (41.97±8.07) |  |
| 30-39 | 184 | 42.9 |
| 40-49 | 158 | 36.8 |
| 50 and above | 87 | 20.3 |
| **Marital status** |  |  |
| Married | 399 | 93 |
| Divorced/widowed/separated | 30 | 7 |
| **Education** |  |  |
| Class 1 to 8 | 247 | 57.6 |
| Class 9 to 12 | 162 | 37.8 |
| Higher than class 12 | 20 | 4.7 |
| **Ethnicity** |  |  |
| Brahmin/Chettri | 174 | 40.6 |
| Janajati | 226 | 52.7 |
| Dalit and others | 29 | 6.8 |

**Table 2.** Prevalence of depressive symptoms, domestic violence and emotional abuse (n=429).

| Prevalence | n (%) | 95% CI |
| --- | --- | --- |
| Depressive symptoms<br>(mean±SD) | 200 (46.6)<br>(11.18±3.43) | 0.42, 0.51 |
| Any type of violence | 54 (12.6) | 0.09, 0.16 |
| Physical violence | 44 (10.3) | 0.07, 0.13 |
| Sexual violence | 8 (1.86) | 0.01, 0.03 |
| Emotional abuse | 33 (7.69) | 0.05, 0.10 |
| Sexual violence or emotional abuse | 36 (8.39) | 0.06, 0.11 |

*The cut off score taken for PHQ-9 scale was 10 i.e., 0 to 9 as absence and 10 and higher as presence of depressive symptoms.*

Almost half of the participants (46.6%) had depressive symptoms. The mean score of depressive symptoms was 11.83 (SD=3.43). Overall, the prevalence of domestic violence was 12.6%. Most of the participants (10.3%) reported that the violence they experienced was physical violence, followed by sexual violence or emotional abuse (8.39%).

Table 3 shows that the most common form of physical violence experienced by the participants was being slapped (11%) while that of emotional abuse was being humiliated in front of others (7.8%). None of the participants reported that they have been threatened or attacked with a knife, gun or other weapon by their partners. Least of them (1.5%) said that they have been burned or choked on purpose. Overall, only one of the participants reported that they were forced by their partners with threats or in any other way to perform unwanted sexual acts.

**Table 3.**
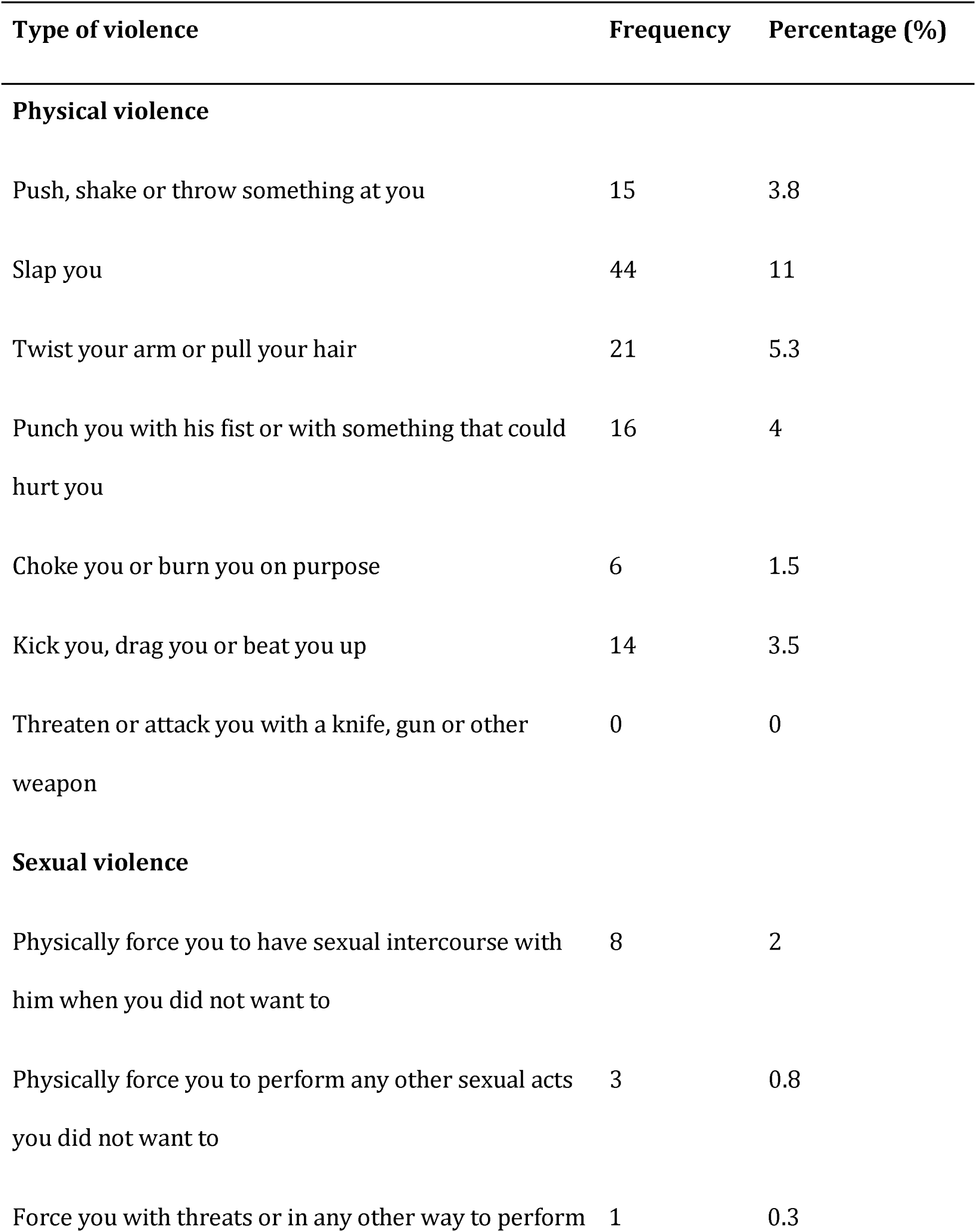
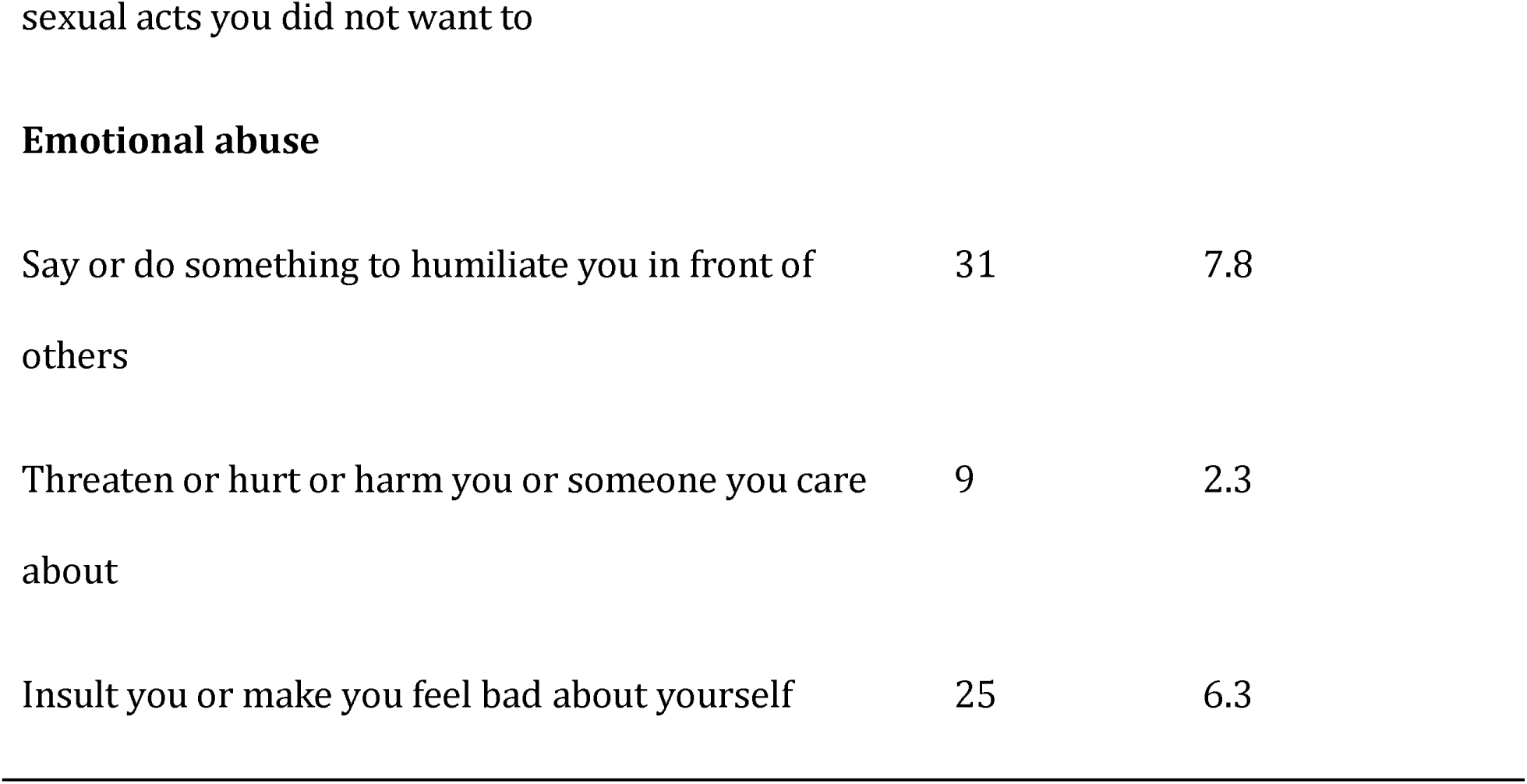
Items used to assess domestic violence (n=429).

Table 4 depicts the crude and adjusted association between violence and depressive symptoms. When we compare two groups of participants with and without experience of any type of violence, the odds of having depressive symptoms were 2.99 times (95% CI: 1.57, 5.67, p = 0.001) higher among those who had experienced any type of violence. Similarly, the odds of having depressive symptoms were 2.5 times (95% CI: 1.26, 4.98, p=0.009) more in participants who had experienced physical violence and 6.21 times (95% CI: 2.49, 15.46, p<0.001) higher among the participants who had experienced sexual violence or emotional abuse.

**Table 4.**
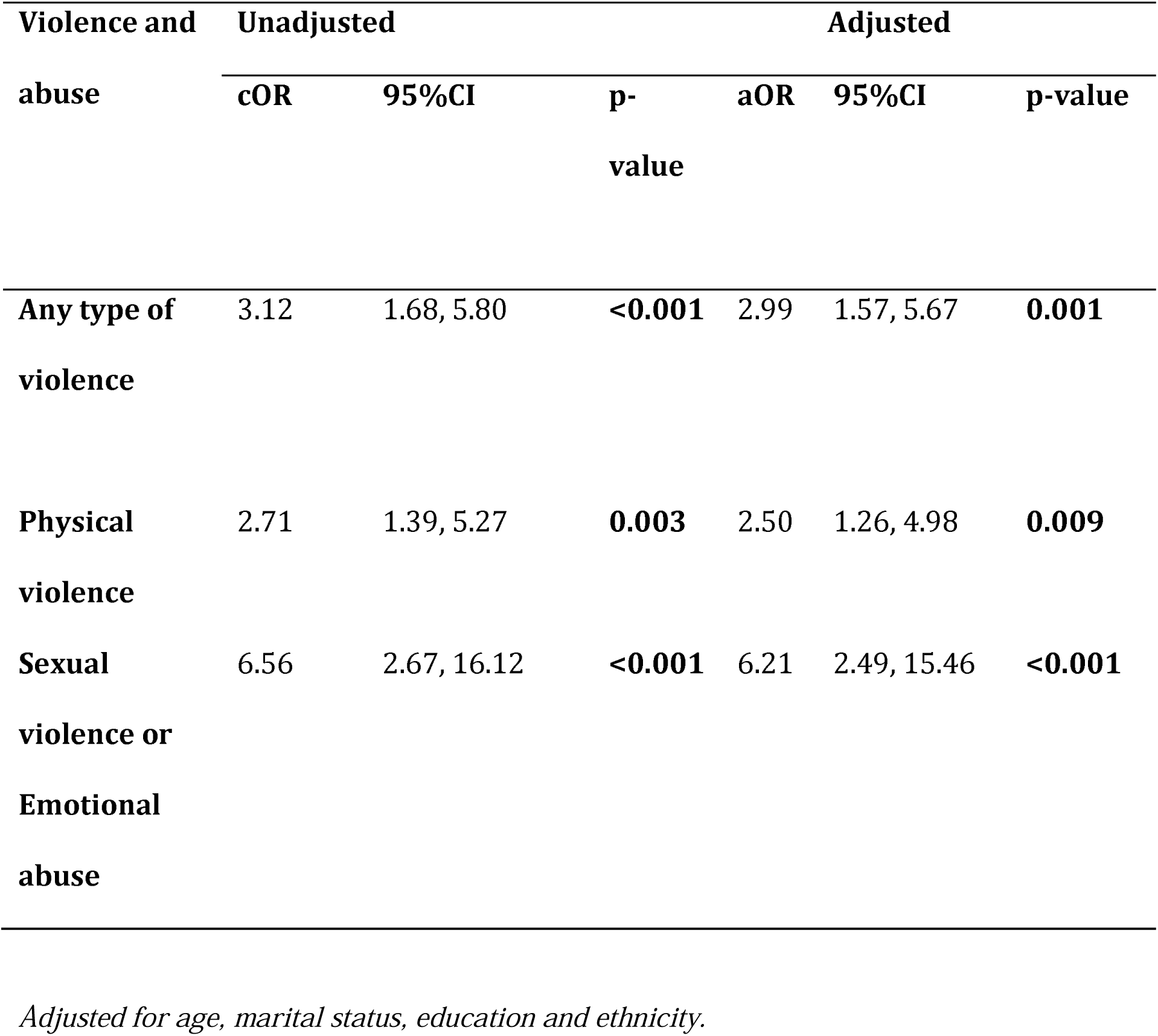
Association of depressive symptoms with domestic violence and abuse (n=429).

| Violence and abuse | Unadjusted |  |  | Adjusted |  |  |
| --- | --- | --- | --- | --- | --- | --- |
|  | cOR | 95%CI | p-value | aOR | 95%CI | p-value |
| Any type of violence | 3.12 | 1.68, 5.80 | <0.001 | 2.99 | 1.57, 5.67 | 0.001 |
| Physical violence | 2.71 | 1.39, 5.27 | 0.003 | 2.50 | 1.26, 4.98 | 0.009 |
| Sexual violence or Emotional abuse | 6.56 | 2.67, 16.12 | <0.001 | 6.21 | 2.49, 15.46 | <0.001 |

*Adjusted for age, marital status, education and ethnicity.*

## Discussion

Findings from our study showed that any type of domestic violence in women is significantly associated with depressive symptoms. In our study, depressive symptoms were prevalent among almost half of the women (46.6%). This finding aligns with existing literature on depression among similar demographics, emphasizing a concerning trend in mental health. For instance, a study conducted in Algeria found that more than half of its participants (58.9%) experienced severe depression (29). Similarly, a study from Indonesia reported a prevalence of severe depression at precisely 50% among its participants (30) while In Pakistan 55.1% of women exposed to IPV reported symptoms of depression (31). This shows depression is a significant issue among women across diverse cultural contexts, despite notable socioeconomic differences between the countries studied.

In this study, 12% of women reported experiencing any form of violence, which is considerably lower compared to the 28% prevalence of any form of intimate partner violence, including physical, sexual, or emotional violence, reported in the national survey of Nepal (NDHS 2022) which was done among women aged 15 to 49 years (32). The NDHS 2022, further indicates that 23% of those women reported physical violence, 12% reported sexual violence, and 7% reported emotional violence, all of which are higher as compared to our study (32). Prevalence of all kinds of violence reported in NDHS is two times or more than that of our study, this may have been affected by the participants’ age range as well. We included women aged 30 to 60 years while NDHS 2022 included women aged 15 to 49 years. Studies have shown that depression and anxiety, like mental health problems affect younger people more than the elder ones (33).

The study conducted in Indonesia with women aged 18 to 60 years showed 9.8% participants experiencing physical violence (30) which is consistent with the findings of our study. However, another study conducted in Algeria with women aged 20 to 60 showed 33.9% prevalence of physical violence among their participants which is remarkably higher (29). Regarding sexual and emotional violence, our study showed that the prevalence of sexual violence or emotional abuse was 8.39%. This is in line with the findings from the Indonesian study where the psychological violence was 8.5% (30). Contrastingly, a study conducted in Indian women showed that the prevalence of psychological violence was much higher (19.8%) (34). These variations in results from different studies might be attributed to differences awareness among women on domestic violence as well as the reporting mechanism of different areas. In areas with limited awareness, women may not recognize certain behaviors as forms of violence, leading to underreporting. In addition to this, there are various stigma, shame, embarrassment and fear attached to the disclosure of the violence.

Our study showed that the women who experienced any type of domestic violence tend to have depressive symptoms 2.99 times more than those who did not experience it. This is supported by the findings of a qualitative study conducted in Pakistani women, in which thematic analysis suggested a close relation of domestic violence with depression (35). Another longitudinal study conducted in the UK showed that there is an association of IPV with depression not only in women but also in men. Women who experienced IPV had double the odds (OR 2.10, 95% CI: 1.57, 2.81) of having depressive symptoms above the threshold defining depression while the men had a 36% higher the odds (OR 1.36, 95% CI: 0.91, 2.04) (36). These findings suggest that violence in general is positively associated with depression irrespective of the individual’s gender. Being a victim of domestic violence has been associated with several physical as well as mental health effects. It has been linked with increased stress, fear, and isolation and eventually to depression and suicidal behaviour or thoughts (37)

Our study showed that women who experienced physical violence had the higher odds of having depressive symptoms. Similar findings are shown by another study done in Sweden, where women who were exposed to physical violence had 3.78 times higher odds of having depressive symptoms (95% CI: 1.99, 7.17) (38). Similarly, in Pakistan, women exposed to intimate partner violence had 1.97 times higher odds of experiencing depressive symptoms (31). In Norway, physical abuse was also significantly associated with depression with an adjusted odds ratio of 2.35 further supporting the association between physical violence and depression (39). The consistency of findings across diverse geographical contexts highlights the global relevance of this association. Cultural and societal norms may influence the prevalence and reporting of violence, but the psychological impact remains with similarity. Violence can leave women feeling helpless, diminish their self-esteem, and lead to social isolation, all of which heighten the risk of depression, a condition closely linked to a history of exposure to stressful life events (40, 41).

Our study also suggested that the women who had ever experienced sexual violence or emotional abuse were likely to have 6.21 times more odds of depressive symptoms compared to those who had not. This is consistent with the findings of the study done in Sweden where women who experienced sexual violence had higher odds of having depressive symptoms (OR 5.10, 95%CI: 1.74, 14.91) (38). In South Africa, past-year exposure to sexual or intimate partner violence was associated with an increased likelihood of depressive symptoms (aOR 2.05) (42). Some studies have suggested that the effects of emotional violence are more severe and even last longer when compared to the effects of physical violence (43). This is due to psychological mechanisms where emotional abuse, rooted in manipulation and control, can create lasting psychological scars and a cycle of revictimization (44). Those effects may include depression, anxiety, panic disorders in addition to a distorted self-image or broken sense of self. Some victims ultimately even admit that they deserved to be abused (13)

This study has several key strengths. First, it is one of the few studies to determine the association between depressive symptoms and the experience of domestic violence and abuse, contributing valuable evidence to a relatively under-researched area. Second, we utilized the PHQ-9, a validated tool for measuring depressive symptoms, with the Nepali version demonstrating adequate internal consistency with the Cronbach’s alpha of 0.84 (28). Lastly, the result of this study are the output of rigorous statistical analysis done after adjusting for potential confounding variables. However, there are several limitations to consider. First, the cross-sectional design prevents us from establishing a temporal relationship between depressive symptoms and experiences of domestic violence. Second, given that the questionnaires were administered by the interviewer, there is a potential for social desirability bias, which may have resulted in underreporting of both depressive symptoms and experiences of violence. Third, the findings may lack generalizability, as the study was limited to women from only two municipalities. Lastly, although we adjusted for known confounders, there is the possibility of residual confounding that could introduce bias into the results.

### Conclusions

Those women who have experienced physical violence, sexual abuse, or emotional abuse are more likely to experience depressive symptoms. To better understand the factors contributing to these symptoms, further in-depth and exploratory studies are needed. Community-based interventions are needed to offer counseling, support groups, and safe spaces for women affected by violence. Additionally, continuous research on the relationship between violence and mental health is crucial to guide policy development and intervention strategies.

## Declarations

### Ethics approval and consent to participate

The ethical approval was obtained from Kathmandu University Institutional Review Committee (KUIRC no: 35/2021). Verbal informed consent was obtained from participants due to telephonic mode of data collection and audio recorded with their consent.

### Consent for publication

Not applicable

### Availability of data and materials

The datasets used and/or analysed during the current study are available from the corresponding author on reasonable request.

### Author’s contributions

AS and BP designed and conceptualized the study. AK and SM had major role in data acquisition. BS and PP analyzed and interpreted the data of the study. BS, YA, PT were major contributors in drafting the manuscript. BS, YA, PT, PP, AK, SM, BP and AS rigorously contributed in editing, revising and proofreading the manuscript. All the authors read and approved the final manuscript.

## Data Availability

All data produced in the present study are available upon reasonable request to the authors.

## Acknowledgements

We are extremely thankful towards Dhulikhel hospital for conducting cervical cancer screening programs, which is the major source of participants for this study.

## List of abbreviations

DSM: Diagnostic and Statistical Manual
FCHV: Female Community Health Volunteer
GBV: Gender-based Violence
IPV: Intimate Partner Violence
NDHS: Nepal Demographic and Health Survey
OCMC: One Stop Crisis Management Center
PHQ: Patient Health Questionnaire
RA: Research Assistant
USAID: United States Agency for International Development

